# Longitudinal evaluation of anti-SARS-CoV-2 neutralizing antibody levels in 3-dose homologous (mRNA-1273- mRNA-1273- BNT162b2) vaccinated Kidney transplant population: 18-month follow-up

**DOI:** 10.1101/2025.02.05.25321720

**Authors:** Ridma P. Karunathilake, A Kumara, A Karunathilaka, AWM Wazil, N Nanayakkara, K Bandara, R Abeysekera, F Noordeen, IB Gawarammana, Champa N. Ratnatunga

## Abstract

**Background:** Kidney transplant recipients (KTRs) were given a 3-dose primary series of COVID-19 vaccination as they were vulnerable to infection due to immunosuppression.

**Methods:** This study was a longitudinal evaluation of nAB dynamics in 43 KTRs in a low-middle income setting receiving 3-dose homologous (mRNA-1273- mRNA-1273- BNT162b2) vaccination against COVID-19. Samples were obtained at time-points (TP) 0- pre-vaccination, TP1- 1 month post first dose(mRNA-1273), TP2-1-month post second dose (mRNA-1273), TP3- 4 months post-second dose, TP4- 2 weeks post-third dose(BNT162b2), TP5-5 months post-third dose and TP6-12 months-post third dose. Anti-SARS-CoV-2 nAB were detected using Genscript cPassTM pseudoviral neutralization kit. Demographic and clinical details were obtained through interviewer administered questionnaires.

**Results:** Pre-vaccination serum analysis showed n=7 KTRs had prior COVID-19 infection, classified as ‘infected+vaccinated,’ while others were ‘vaccinated. ‘ Both groups were similar in age(41.7years vs 46.7years,*p=0.2383*), gender, and transplant characteristics. Seroconversion and MAB in the vaccinated and infected+vaccinated KTRs were:TP1-8.3% vs 100%(*p<0.001*), MAB-64.3IU/ml vs 1424IU/ml(*p=0.0167*TP2-52.7% vs 100%(*p=0.0194*), MAB-175IU/ml vs 2790IU/ml(*p<0.0001*), TP3-100% vs 100%, MAB-106IU/ml vs 2153IU/ml(p=0.0002), TP4-100% vs 100%, MAB-736 IU/ml vs 2152IU/ml(*p=0.0307*) and TP6-100% vs 100%, MAB >2565IU/ml vs >3028IU/ml(*p=0.5238*) No factors were associated with seroconversion or MAB.

**Conclusion:** KTRs receiving a three-dose mRNA COVID-19 vaccine regime maintained strong nAB levels at one-year follow-up, with comparable antibody levels seen between KTRs with prior infection + vaccination and vaccination alone.

## Introduction

Kidney transplant recipients (KTRs) hospitalized in the early stages of the COVID-19 pandemic faced serious health consequences. Early data showed high (20%-30%) SARS COV-2 infection rates (1,2) while the absolute number of deaths during the first wave was over two folds greater than that seen in previous years. Of these, 44% were due to COVID-19(3). Compared to the average global infection fatality rate of 10.73 per 10,000 infections, (4) KTRs displayed a high mortality rate of 20%-30%(5,6,15,16,7–14)

Immunosuppression mediated vulnerability to both primary pathogenic and opportunistic infections is a well know hazard of being a transplant recipient. Poor immunity also equates to longer pathogen clearance times, increasing the risks associated with prolonged harbouring of a virus, which include in-host viral mutations and being a prolonged source of transmission(17). Priority SARS-CoV-2 vaccination for KTRs was therefore recommended and practiced globally (18).

Weak immune responses to other vaccines including the trivalent influenza vaccine in KTRs on mycophenolate mofetil therapy raised a reasonable doubt whether an initial two-dose primary series of COVID-19 vaccination would be sufficient to provide protection from infection/ severe disease(19). Confirming these suspicions, early studies revealed that compared to the high seroconversion rate in healthy adults, only 22%-36.4% of KTRs developed anti-SARS-CoV-2 IgG antibodies after 2 doses of BNT162b2(16)(20) and only 48%-65% after 2 doses of mRNA- 1273(21)(22). Even when formed, antibodies waned rapidly over a 6 month period(23). Given the low antibody levels and almost absent T cell responses seen in seroconverted KTRs(24), additional vaccine doses for these vulnerable individuals was recommended.

The interim guidelines for COVID-19 vaccination issued by the Strategic Advisory Group of Experts (SAGE) of the World Health Organization (WHO) in October 2021, recommended that all immune compromised patients receive a three-dose primary series where possible(25). Recent studies confirmed the improved response to third dose of mRNA vaccine in achieving sustained seroconversion and antibody levels in KTRs(26–28).

Our study focused on a cohort of KTRs in Sri Lanka, a lower middle-income country that went through four waves of COVID-19 from March 2020 to December 2022, with consequent economic crisis and social upheaval, severely affecting all aspects of life. However, the national vaccine programme provided priority vaccination for patients on dialysis and KTRs from July 2021. COVID-19 related mortality in this patient group was 33.3% prior to vaccination in Sri Lanka (29). Patients were given a three-dose homologous vaccine combination of mRNA-1273, mRNA-1273, BNT162b2, based on in-country vaccine availability. We evaluated the longitudinal neutralizing antibody (nAB) dynamics throughout the completion of the vaccination and continued a one-year follow-up after third dose while analysing the factors associated with seroconversion and identifying vaccine associated adverse events. The objective was to study the longevity and level of neutralizing humoral responses in these patients.

## Materials and Methods

### Study population

This longitudinal study was carried out between July 2021 and December 2022, up to 1 year following vaccine third dose administration to KTRs. Initially, 43 KTRs who attend the Nephrology unit of National Hospital Kandy, the largest kidney transplant centre in the country, were recruited. Ethical clearance (EC/2021/M/14) was obtained from the Ethics Review board of the Faculty of Medicine, University of Peradeniya, Sri Lanka.

The KTRs were given two doses of mRNA-1273 one month apart, and a third vaccine dose of BNT162b2, 4 months later, to complete the primary series. Serum sample collection was done at seven time points in relation to vaccine administration as shown in **Fig 1**; Pre vaccination (TP0), 1month post first dose (TP1), 1 month post second dose (TP2), 4 months post second dose (TP3), 2 weeks post third dose (TP4), 5 months post third dose (TP5) and 1 year post third dose (TP6).

**Figure 1.** Timeline of vaccination and sampling time points. The timeline shows the timing of vaccination and sampling time points of the patient cohort along with timing of concurrent waves of COVID-19.

Sociodemographic data, history of symptoms suggestive of COVID-19 infection or COVID-19 confirmed by RT-PCR/ antigen detection (testing was freely available at the study site throughout period of study), and vaccine adverse events related to each vaccine dose were collected from each participant at each time point using an interviewer administered questionnaire. Furthermore, duration of transplant, history of transplant rejection and patients on immunosuppressive drugs (prednisolone, mycophenolate mofetil (MMF), tacrolimus, cyclosporin, everolimus) were documented.

Over the period of follow-up, from TP2 to TP3 sample numbers had to be restricted due to limited test availability, while during subsequent timepoints, dropouts due to refusal or inability to participate because of logistic/ economic reasons were noted. A summary of participant recruitment and follow-up is shown in **S1 fig.** When sampling restriction was required, participants who agreed to continued participation for long term testing, planned to take the third dose, and were readily contactable, were selected.

Blood samples were obtained to detect the presence of neutralizing antibodies against SARS-CoV-2 using the GenscriptC Pass^TM^ SARS-CoV-2 neutralization Antibody Detection kit (GenScript USA/ Nanjing GenScript Diagnostics Technology Co., Ltd., Version RUO for US.1.0). Separated serum samples were stored at −80°C until testing. Testing was carried out according to manufacturer’s instructions. Percentage inhibition (calculated based on manufacturer instructions) was obtained using the surrogate viral neutralization assay. Seroconversion was defined as percentage inhibition value above a cutoff of 30% neutralization, while level of antibody was equated to the percent neutralization calculated. An additional measure of antibody level was calculated based on WHO guidelines(30) and conversion algorithm published for this assay(31). Percentage neutralization value for each sample was converted into IU/ml based on reference algorithm. Values above 97.59% neutralization (equivalent to >3028 IU/ml) required further dilution and re-testing for accurate conversion to IU. Dilution retesting was done in only 8/18 samples (from TP1 and TP2) that required retesting due to financial constraints. Samples requiring dilution testing from TP6 in both groups (‘vaccinated’ n=8 and ‘infected + vaccinated’ n=2) could not be retested. For visualization, these samples values are represented as >3028 IU/ml in plots and samples that could not be retested were included as 3028 IU/ml in statistical testing. The conversion algorithm was log scaled; therefore, mean values are expressed as geometric mean with a SD factor. Results are therefore expressed as seroconversion percentage (the number of individuals with antibody levels above the defined cut-off), mean/ median antibody level (MAB - the midpoint estimate of percentage neutralization of the positive samples within the group).and the equivalent geometric mean antibody unit level as IU/ml.

### Statistical analysis

Statistical analysis was carried out using GraphPad Prism (8.4.2. 679, 2020) Percentage neutralization was compared using Mann Whitney test and association between categorical variables was evaluated using the chi-square test(32).

## Results

Initially, n=43 KTRs were recruited in whom pre vaccination samples (TP0) revealed seven (7/43) with positive neutralizing antibodies despite having no history of suggestive symptoms or RT-PCR test positivity. This patient subset was categorized as ‘infected + vaccinated’ KTRs. The pre-vaccination seronegative KRTs (n=36) were categorized as ‘vaccinated’ KTRs. During follow up none of the participants developed clinical symptoms suggestive of, or RT-PCR confirmed SARS-CoV-2 infection. Number of serum samples in each group at each timepoint are shown in **S1 fig**.

### Demographic characteristics

#### Age and gender distribution

Vaccinated KTRs and infected + vaccinated KTRs were similar in age (41.7 years (SD+/-11.2)) vs 46.7 years (SD+/-11.2, MWU, p=0.2383), as well as gender distribution (chi sq, df2, p=0.8429) with both groups having a male majority. Duration of transplant, history of transplant rejection and the number of KTRs on different immunosuppressive drugs are shown in **Table 1**. Mean duration of transplant in the vaccinated KTRs (56.6 months) was similar (MWU, p=0.3108) to that of infected + vaccinated KTRs (63.5 months) with an overall range of 10 months to over 15 years while nearly half of the total KTRs had undergone kidney transplant, 2-5 years ago. Prednisolone (vaccinated:100%, infected+vaccinated:100%), MMF (vaccinated: 97.6%, infected + vaccinated: 85.7%, chi sq, p=0.1628) and Tacrolimus (vaccinated: 94.4%, infected + vaccinated:100%, chi sq, p>0.9999) were the most common immunosuppressive drugs that the patients in both groups were on, in similar frequency. A summary of immunosuppressant medication doses and patient numbers are provided in **S2 Table**. KTRs in both groups had been receiving a maximum of three immunosuppressive agents. History of transplant rejection was similar in both groups (vaccinated: 63.8% (23/36), infected + vaccinated :71.4% (5/7), chi sq,p=0.7017). All the KTRs were vaccinated with three-dose Hepatitis B vaccine series though the seroconversion status is unconfirmed.

**Table 1.** Kidney transplant recipient characteristics.

| Characteristic | Value/ Frequency |  | P value |
| --- | --- | --- | --- |
|  | Vaccinated | Infected + Vaccinated |  |
| Age |  |  |  |
| Mean (SD)/median (IQR) | 41.7 (11.2)years/<br>40.5 (32.5- 50) | 46.7 (11.2) years<br>50 (39- 55) | p=0.2383<sup>b</sup> |
| Range | 23-69 years | 26-60 years |  |
| Gender |  |  |  |
| Male | 75% (27 /36) | 71.4% (5 /7) | p=0.8429<sup>a</sup> |
| Female | 25% (9 /36) | 28.6% (2 /7) |  |
| Transplant duration |  |  |  |
| Mean (SD)/median (IQR) | 56.6 (43) months/<br>48.5 (26-77) | 63.5 (35) months/<br>57 (36-77) | P=0.3108<sup>b</sup> |
| Range | 10-190 months | 32-132 months |  |
| <2 years | 22.2% (8 /36) | 0 |  |
| 2-5 years | 41.6% (15 /36) | 57.2% (4 /7) |  |
| 5-10 years | 25% (9 /36) | 28.6% (2 /7) |  |
| > 10 years | 11.2% (4 /36) | 14.2% (1 /7) |  |
| Patients with a history of transplant rejection | 63.8% (23 /36) | 71.4% (5 /7) | p=0.7017<sup>a</sup> |
| Patients on immunosuppressive drugs |  |  |  |
| Prednisolone | 100% (36 /36) | 100% (7 /7) |  |
| MMF | 100% (36 /36) | 85.7% (6 /7) | p=0.1628<sup>a</sup> |
| Tacrolimus | 94.4% (34 /36) | 100% (7 /7) | p>0.9999<sup>a</sup> |
| Cyclosporin | 2.7% (1 /36) | 0 |  |
| Everolimus | 11.1% (4 /36) | 14.2% (1 /7) | p>0.9999<sup>a</sup> |

<sup>a</sup>Chi sq/ Fisher’s exact test <sup>b</sup>Mann-Whitney U test (MWU)

### Longitudinal Antibody dynamics following two doses of mRNA-1273 and a third dose with BNT162b2 in vaccinated KTRs

Neutralizing antibody dynamics in the both KTR groups (vaccinated and infected + vaccinated) following two doses of mRNA-1273 and a third dose with BNT162b2 are shown in **Fig 2**. All patients in the ‘vaccinated’ group were seronegative prior to vaccination. The percentage seropositive at TP1, one month after the first dose was 8.3% (03/36) with a moderate median antibody level (MAB) of 54.85% neutralization (IQR-32.59%-59.49%) which is correspondent to 64.37 IU/ml (SD factor-1.84). At TP2, 2-4 weeks following the second dose, seropositivity significantly increased (chi sq, p<0.001) to 52.7% (19/36) whilst MAB increased (MWU, p=0.1325) to 72.90% neutralization (IQR:44.70%-86.84%)/ correlating to a unit value of 175.40 IU/ml (SD factor-3.09). At TP3, four months following second dose, seropositivity significantly increased again (chi sq, p<0.001) to 100% (21/21) though MAB dropped somewhat (MWU, P=0.2055) to 51.96% neutralization (IQR:39.84%-80.90%)/ with a unit value 106.6 IU/ml (SD factor-3.25). The changes in antibody levels (MAB and unit value as IU/ml) were not statistically different between any of these initial timepoints. However, a deeper look revealed that while all vaccinated KTRs had become seropositive by TP3 (21/21), the 52.4% who were seronegative at TP2 (11/21) showed significant increase in nAB levels at TP3 (mean % inhibition/IU/ml (95%CI): 21%(18.4-24.3%)/17.5 IU/ml(14.6-20.9 IU/ml) vs 41.2%(35.1%-47.3%)/46.7 IU/ml (36-60.4 IU/ml), paired t test, p<0.0001), while the 47.6% who were seropositive at TP2 (10/21) showed no significant difference in antibody levels (70.5% (56.6-84.3%)/ 195 IU/ml(93.8-404.7 IU/ml) vs 74.7% (62.6-86.9%)/ 264.6 IU/ml(122-574 IU/ml), paired t test, p=0.5488))) at TP3.

**Figure 2.** Longitudinal neutralizing antibody (nAB) dynamics following two doses of mRNA-1273 and a third dose of BNT162b2. Dot plot shows neutralizing antibody dynamics in KTRs with natural infection + vaccination (orange circles) and vaccine only (open circles) at the time points of pre-vaccination (TP0), 1 month post first dose (TP1), 2 weeks post second dose (TP2), 4 months post second dose (TP3), 2 weeks post third dose (TP4), 5 months post third dose (TP5) and 12 months post third dose (TP6). Primary Y axis (blue) shows antibody level in IU/ml, while secondary y axis (black) shows the equivalent percentage neutralization value. Seropositivity designated at >30% (28 IU/ml) (manufacturers standard). Data points plotted at >3028 IU/ml (>97.59% inhibition) indicate the individuals with antibody levels that range between 3878-29133 IU/ml. A significant increase in antibody levels in ‘vaccinated KTRs’ was seen after third dose vaccination (TP3 to TP4) and during the one-year follow-up period (TP4 to TP5; TP5 to TP6). Infection + vaccinated KTRs showed persistently high nAB levels throughout follow up. Red dashed lines indicate the time points of vaccine administration. (D1– first dose, D2– second dose, D3-third dose). Statistically significant difference in MAB (Mann Whitney U test) denoted by p<0.05-*, p<0.01-**, p<0.001 - ***, p<0.0001 - ****.

Two weeks following the administration of the BNT162b2 vaccine as the third dose (TP4), seropositivity remained constant at 100% (21/21) while the MAB significantly increased (MWU, p<0.0001) to 95.08% neutralization (IQR:85.88%-96.05%)/ with equivalent unit value of 736.50 IU/ml (SD factor-3.58). At TP5, 5 months after dose 3, seropositivity significantly reduced (chi sq, p<0.05) to 81.25% (13/16) though the MAB of the seropositive KTRs increased significantly (MWU, p=0.0065) to 97.06 % neutralization (IQR:88.74%-97.29%)/ 1459 IU/ml (SD factor-2.58). A deeper analysis showed that 76.9% (10/13) of KTRs who were seropositive at TP5 showed increasing nAB levels compared to TP4 ((95.7% (IQR:88.7-96.3%)/1062 IU/ml (595-1896 IU/ml) vs 97.2% (IQR:96.9-97.3%)/ 2363 IU/ml (1973-2829 IU/ml), Wilcoxon test, p=0.0020)), while 23% (3/13, seropositive at TP5) showed a decreasing trend without a significant difference in nAB levels((94% (IQR :91.8-95%)/1095 IU/ml (544.6-2200 IU/ml) vs 82.4% (75.4-83.2%)/292.6 IU/ml (150-570 IU/ml, Wilcoxon test, p=0.2500). Three KTRs (18.7% of the total tested at TP5) sero-reverted at 5 months post third dose. As shown in **Fig 2**, these were KTRs who showed persistently low antibody levels. despite late seroconversion four months after second dose. Other KTRs who showed late seroconversion after second dose (n=8) showed significant boost in antibody levels following the third dose. There was no significant difference in terms of age, gender, duration of transplant, type of immunosuppressant or history of transplant rejection between the KTRs who had persistently low nAB responses, and subsequently sero-reverted, compared to the other low responders who later showed good responses to the third dose.

Twelve months following the third dose (TP6) 100% (13/13) seropositivity was observed, while MAB significantly increased (MWU, p=0.0075) up to 97.85% neutralization (IQR:96.77%-98.01%)/ with a very high equivalent unit value of 2565 IU/ml (SD factor-1.35). While 61.5% (n=8) of patients showed a consistent increase in antibody levels with, 15% (n=2) of patients who showed an initial drop at 5 months also showed a subsequent increase at 1 year follow-up. A lack of one year data on the KTRs who seroreverted at 5-months is noted here. A comparison of % seroconverted and MAB level (geometric mean IU/ml and % inhibition) between each consecutive time point in the vaccinated KTRs is shown in **S3 Table.**

#### Longitudinal antibody dynamics in ‘Infected + vaccinated’ KTRs

Prior to vaccination (TP0), seven KTRs found to be positive for nAB had a median antibody level of 93.39% neutralization (IQR:39.61%-96.85%)/ which is equivalent to a unit value of 390 IU/ml (SD factor-7.11). This initial MAB did not exhibit a significant change after the first dose (TP1-95.12%, IQR:94.02%-97.74% / 1424 IU/ml, SD factor:2.66, MWU, p=0.1649) or second dose at either TP2 (97.71%, IQR:97.02%-97.75% / 2790 IU/ml, SD factor: 1.15, MWU, p=0.1183) or TP3 (96.82%, IQR: 95.51%-97.38% / 2153 IU/ml, SD factor:1.35, MWU, p=0.0727) though an increasing trend was seen. Range of nAB in dilution tested samples was 3878 IU/ml – 29133 IU/ml. Sample collection following the third dose was low in number, however, collected samples revealed no significant change in nAB levels following third dose by TP4 (96.66%, IQR: 95.7%-97.4% / 2152 IU/ml, SD factor:1.32, MWU, p=0.8857). While logistic difficulties prevented sample collection at TP5, sustained high antibody levels being seen at one year follow-up (TP6 - >97.98%, IQR: 97.92%-98.04% / >3028 IU/ml, SD factor:1, MWU,p=0.1333), where both samples collected had Nab levels requiring dilution retesting.

### Comparison between ‘vaccinated’ group and ‘infected + vaccinated’ group

The MAB and seroconversion rate in KTRs with known natural infection + vaccination and vaccinated KTRs at each time point were compared (**Table 2**). The naturally infected KTRs maintained 100 % seroconversion throughout the course of follow-up, which was significantly higher than that of the vaccinated KTRs group at TP1 and TP2. nAB levels were also consistently higher compared to the infection naïve group throughout the period starting from 1 month post first dose (TP1) to 2 weeks post third dose (TP4). Both groups showed very high MAB at 1 year post third dose. Statistical comparison could not be done at TP5 due to lack of samples in ‘infected + vaccinated’ group.

**Table 2.** Comparison between ‘vaccinated’ KTRs and ‘infected + vaccinated’ KTRs - percentage seroconversion and MAB.

|  |  | D1 | D2 | D3 |  |  |
| --- | --- | --- | --- | --- | --- | --- |
| Measure |  | TP1<br>1 mo post<br>first dose | TP2<br>1 mo post<br>second dose | TP3<br>4 mo post<br>second dose | TP4<br>2 wks post<br>third dose | TP6<br>12 mo post<br>third dose |
| Infected +<br>Vaccinated<br>KTRs | % Seroconverted | 100% | 100% | 100% | 100% | 100% |
|  | <sup>a</sup>MAB geo.mean<br>IU/ml<br>(%inhibition) | 1424<br>(95.12%) | 2790<br>(97.71%) | 2153<br>(96.82%) | 2152<br>(96.66%) | 3028<br>(97.98%) |
| Vaccinated<br>KTRs | % Seroconverted | 8.3% | 52.7% | 100% | 100% | 100% |
|  | <sup>a</sup>MAB geo.mean<br>IU/ml<br>(%inhibition) | 64.37<br>(54.85%) | 175.4<br>(72.90%) | 106.6<br>(51.96%) | 736.5<br>(95.08%) | 2565<br>(97.85%) |
| *P* value | <sup>b</sup>*Chi sq* | *p*<0.0001 | *p*=0.0194 | *p*>0.9999 | *p*>0.9999 | *p*>0.9999 |
|  | <sup>c</sup>*MWU* | *p*=0.0167 | *p* < 0.0001 | *p*=0.0002 | *p* =0.0307 | *p*=0.5238 |
|  | (*MWU*) | (*p*=0.0167) | (*p* < 0.0001) | (*p*=0.0002) | (*p* =0.0307) | (*p*=0.2286) |

<sup>a</sup>MAB- mean/ median antibody level of seropositive at each time point. Represented as geometric mean (geo.mean) in International Units (IU/ml) and as mean percentage inhibition (% inhibition).
<sup>b</sup>Chi sq test was performed to compare the % seroconverted between vaccinated and infected+ vaccinated groups at each time point.
<sup>c</sup>Mann Whitney U (MWU) test was performed to compare the MAB levels of seropositives between vaccinated and infected+ vaccinated groups at each time point in both international units (IU/ml). Comparison of % inhibition between each consecutive timepoint is shown within brackets. (Fishers exact test used when sample size <5). Significance at *p*<0.05

### Factors associated with seroconversion and antibody level

None of the demographic or clinical factors such as duration from the kidney transplant, history of transplant rejection or type of immunosuppressive therapy were associated with seropositivity or neutralizing antibody level in this study.

### Vaccine associated adverse events (VAAE) after each dose of mRNA-1273

After the first dose, more than two thirds of patients experienced pain at the injection site (79%) while the least common side effects were headache (6.98%), arthralgia (6.98%) and nausea (4.65%). Fatigue, myalgia and fever were experienced by 25.6%, 20.9% and 13.9% of the cohort respectively. The most common side effect following the second dose was also the pain at the injection site (74.4%) while less than 15% of the cohort experienced each of the fever (11.6%), headache (11.6%), arthralgia (7%), malaise (7%), nausea (4.6%) and chills (4.6%). Fatigue and myalgia were experienced by 37.2% and 32.5% of the KTRs respectively. None of the participants experienced VAAEs after the third dose.

## Discussion

In this study we demonstrate the immune response in terms of nAB dynamics to homologous 3-dose SARS CoV-2 mRNA vaccine regime (mRNA-1273-mRNA-1273-BNT162b2) in a cohort of KTRs over a period of 18 months, up to 1 year post third dose with no further boosters. To the best of our knowledge this is the first study on nAB levels following 3-dose mRNA vaccination providing 1 year follow-up data in KTRs.

We show a seroconversion rate of 52.7% with a MAB of 175.4 IU/ml after two doses of mRNA-1273, in infection naïve KTRs, similar to the seroconversion (42% −58%) and MAB (25 BAU/ml- 142.1 U/ml) observed in several other studies on solid organ transplant and KTRs (2 doses mRNA vaccine) (33)(34). A systematic review and meta-analysis of 44 studies involving 5892 KTRs indicated the overall seroconversion rate (the % of nAB, anti-SARS-CoV-2 spike RBD, or either IgG or IgA anti-spike protein that indicated above-normal quantification results) following 2 doses of mRNA-1273 vaccine in KTRs was 39.2%(35). Seroconversion rates calculated will vary with the type of assay used (anti S1 IgG/ IgA vs nAB etc.), assay type and its inherent detection limit, and positivity cut-off, as well as the time post second dose that sampling was performed as antibody response patterns are not uniform in these immune suppressed individuals. Slow humoral responses and delayed seroconversion is a known phenomenon in transplant recipients(36).

A large study with six-month follow-up of 2 dose vaccinated KTRs showed that by six months, of the 57% of anti-receptor binding domain (RBD) positive patients, 54.8% had decreasing, and 33.7% had equal antibody levels compared to the two month timepoint, while 11.5% had increasing levels, and 5.8% showed de novo seroconversion(37). This phenomenon was seen only in KTRs and not in other patients with kidney disease who received vaccination. Similar findings for influenza vaccination(38) and COVID-19(39) seem to indicate that the immune suppression used was causing a delayed humoral response as well as a decline in antibodies, though the specific mechanism or culprit drug/s are yet to be elucidated. Our data, shows a similar phenomenon with 52.4% of the seropositive KTRs at four months post second dose showing delayed seroconversion possibly due to de novo seroconversion or further exposure to the virus during the follow-up period.

The lack of cross neutralizing antibodies providing protection from variants, as well as very poor T cell responses after two-dose vaccination has also been shown(36)(37). These findings provide the basis for the persistent risk of severe disease and high mortality (5-10%) in KTRs who received only two vaccine doses(40)(41).

Third dose administration, six months following dose two was advocated and adequate vaccines were made available. However, vaccine uptake in the community was at very low levels due to travel costs, fuel shortages and other consequences of a severe economic downturn. Only 21/43 (48.8%) of the originally recruited KTRs received dose three in this study group. We show a significant increase in nAB levels following third dose administration which is consistent with global data showing significant improvement of seroconversion and MAB after the third dose of 3-dose homologous /heterologous COVID-19 mRNA vaccine schedule(28)(27)(26) with 27% of the seropositives achieving anti-spike antibody levels more than 4160 AU/ml while that value was as high as 92.9% in healthy controls(27).

At five months post third dose, seropositivity reduced as a similar pattern of a reduction in antibody levels and sero-reversion in 18.7% of patients was seen. This was seen in three patients who showed late seroconversion following dose two, with persistently low nAB levels which subsequently fell below positivity cutoff. However overall, a significant increase in antibody level was noted, a trend which continued at one year follow-up. Other studies on three-dose vaccinated solid organ transplant patients(36) have shown similar excellent responses to third dose vaccination (73%) with antibody levels reaching healthy control comparable levels, with better, albeit suboptimal neutralization of variants. KTR specific data shows 49.5% of post second dose seronegatives developing de novo seroconversion approximately 1 month after dose three (42). Our data shows that increasing antibody concentrations over long periods appears possible, perhaps up to one year though whether this indicates very slow antibody response to vaccine over a long period of time or response to environmental exposure cannot be teased apart in this data set. From approximately December 2021 to April 2022 (4^th^ wave of COVID-19), SARS CoV-2 Omicron BA.5 spread in the community. Sampling at TP5 and TP6 were carried out in the months following this wave. None of the cohort were symptomatic or had RT-PCR, or rapid antigen positive test results during this time. Anti-nucleocapsid antibody testing was not done due to funding limitations, though even this test has shown sensitivity with low seroconversion rates in vaccinated individuals with confirmed infection(43).

It is noteworthy that studies that have evaluated a further 4^th^ dose show that while some patients (20-25%) do not seroconvert even after a third or fourth dose, for those who do seroconvert, antibody levels remain lower than those who responded to two doses(42).

Vaccination in those with previous SARS CoV-2 infection, despite all infections being asymptomatic in this cohort, resulted in higher levels (2790 IU/ml) of immunity after just two doses of COVID-19 vaccination and the nAB levels was equivalent to that acquired by infection naïve group after three vaccine doses (2565 IU/ml). This is similar to the data reported by a UK study which indicate that KTRs with a history of SARS-CoV-2 infection show significantly higher anti-S IgG compared with infection-naïve KTRs following each vaccine of 4-dose COVID-19 vaccine series(42).

In terms of variant neutralization capability and T cell responses, it’s been shown that 100% of the seroconverted KTRs after two doses of COVID-19 vaccines had the neutralization capacity against B.1.1.7 with a median serum dilution that reduced infection of cells by 50% while only 64%(23/36) and 67%(24/36) showed neutralization against B.1.351 and B.1.617.2, respectively(44). Moreover, a fourth mRNA-1273 dose improves nABs against Delta variant from 16% to 66%, in those with a weak third dose antibody response (45).

A systematic review of 37 studies with 3429 KTRs showed de novo seroconversion after third and fourth vaccine doses ranged from 32 to 60% and 25 to 37% respectively while variant-specific neutralization ranged between 59% and 70% for Delta, 12% and 52% for Omicron. (46). These evidence highlights the possibility of at least partial protection from SARS CoV-2 variants in KTRs upon completion of three doses (even the KTRs who showed delayed humoral response) with the neutralization capacity being increased with subsequent booster doses. The longevity of these responses is not yet known as most studies are based on sampling done within 1-3 months post vaccination. The importance of our data, though small in scale, is with respect to showing the long-term maintenance of nAB levels in these vulnerable patients, particularly in a resource limited setting.

In this cohort, none of the demographic or clinical factors were associated with seroconversion including time post transplantation, history of rejection and immunosuppressant dose at any timepoint. However, a meta-regression revealed that low antibody response is significantly associated with high dose mycophenolate mofetil (MMF)/mycophenolic acid, belatacept, and anti-CD25 induction therapy use. In contrast, the use of tacrolimus was associated with higher antibody response(35). Similarly for third dose vaccination, older age, lower eGFR(26), timing of first vaccination after transplantation, immunosuppression burden, and a diagnosis of diabetes(42), were independent risk factors for non-seroconversion while for fourth dose, receiving ≥2 different classes of immunosuppression medications was associated with non-seroconversion(42).

Overall, our study shows that 3-dose homologous primary series of vaccination (mRNA-1273-mRNA-1273-BNT162b2) elicits a robust and durable neutralizing antibody mediated immune response in most KTRs who were available for follow-up. As nAB titres prior to infection is shown to be predictive of protection from day 40 of infection(47) immunity generated either by vaccination or natural infection with vaccination in KTRs who received 3-dose COVID-19 vaccination provides comparable long term protection which could last up to 1 year-post third dose.

An immunocompetent control group who received the same vaccination regime could not be recruited at the time for comparison, so nAB levels in healthy individuals with the same vaccine regimen in this setting are not known. However, a parallel study in the same setting, involving patients on haemodialysis(HD) and its control cohort(HC) though on a different primary vaccination schedule (AZD 1222-AZD 1222-BNT162b2) provides a comparative immune response pattern(48). In this cohort, two weeks after the initial two doses, KTRs had lower MAB (175 IU/ml) compared to the HD group (450 IU/ml, MWU p=0.014)) and the HC group (1940 IU/ml, MWU, p<0.0001). Four months later, KTRs continued to exhibit lower antibody levels (MAB:106 IU/ml) than HD patients (235 IU/ml, MWU, p=0.032) and HCs (453 IU/ml, MWU, p<0.0001). In contrast, following the third dose, the MAB of KTRs (736 IU/ml) became comparable to those in the HD group (1029 IU/ml) and HC (1538 IU/ml). These elevated antibody levels were sustained up to one year with KTRs maintaining (>2565 IU/ml) similar to HD patients (>1961 IU/ml) and HCs (>2911 IU/ml).

Loss to follow-up rates were high, particularly during the peak of the third wave of COVID-19 pandemic in Sri Lanka. Re-testing of samples requiring dilutions was performed in only 8 of the 18 individuals with high % neutralization giving antibody levels ranging from 3878 IU/ml to 29133 IU/ml, 10/18 samples were from ‘infected + vaccinated’ group (TP1, TP2,) while 8/18 were from both groups at TP6. The true values of MAB levels at TP6 are therefore probably much higher than estimated in these calculations. The corollary of this is that better cross neutralization of variants and protection from severe infection is likely.

## Conclusions

KTRs receiving a three-dose (mRNA-1273- mRNA-1273 -BNT162b2) mRNA vaccine regime show high nAB responses at one year follow-up, with comparable antibody levels seen between KTRs with prior infection + vaccination and those who had only vaccination. nAB levels shown to be associated with at least partial variant protection lasting at least one year after third dose indicate robust protection in this patient group.

## Data Availability

All relevant data are within the manuscript and its Supporting Information files.

## Acknowledgement

We extend our gratitude to all the study participants who got involved in the study despite difficult times, Mrs. H.M. Ashoka Shanthi, staff of the Nephrology clinic of National Hospital Kandy for their unwavering support, and the hospital management. We are also grateful to the funding bodies, the University of Peradeniya and Peradeniya Kidney Protection Society for partial funding of this project.

---

**S1 Figure. Participant recruitment in KT cohort.** The flow diagram shows the number of participants recruited, excluded and lost to follow-up at each time point.

**S2 Table. A summary of immunosuppressant medication doses and patient numbers**

**S3 Table. Comparison of percentage seroconverted and mean/ median antibody levels between consecutive time points in Vaccinated KTRs**

**S4 Table. Neutralizing antibody levels of KTRs at each timepoint**

**S5 Table. Kidney transplant recipient characteristics**

## Notes

### Competing Interest Statement

The authors have declared no competing interest.

### Funding Statement

Yes

### Author Declarations

Informed written consent was obtained from the participants. Ethics approval was obtained from the Ethics Review Committee, Faculty of Medicine, University of Peradeniya (2021/EC/14). All methods were performed in accordance with the relevant guidelines and regulations.

